# GPT-4 Enhances Readability of Online Patient Materials for Cardiac Amyloidosis

**DOI:** 10.1101/2025.05.19.25325053

**Authors:** Joseph Haquang, Vishnu Bharani, Jamil S. Samaan, Ryan C. King, Samuel Margolis, Nitin Srinivasan, Nithya D. Rajeev, Joshua Chan, Yee Hui Yeo, Roxana Ghashghaei

**Author notes:** Corresponding author: Joseph Haquang. These authors contributed equally to this work and share first authorship (Joseph Haquang, Vishnu Bharani). 333 The City Blvd W, Suite 400, Orange, CA 92868. **Financial Support:** No external financial support.

## Abstract

**Background:** Cardiac amyloidosis (CA) is a rare, progressive condition that requires complex management, highlighting the importance of accessible and understandable patient education materials (PEMs). The American Medical Association (AMA) recommends that PEMs be written at or below a 6th-grade reading level; however, PEMs for CA often exceed this recommendation. GPT-4, a large language model (LLM), is increasingly being studied for its ability to enhance PEM readability.

**Materials and Methods:** Institutional PEMs were sourced from the websites of the top 10 institutions ranked by the 2022-2023 US News & World Report for “Best Hospitals for Cardiology, Heart & Vascular Surgery.” GPT-4 (version updated 20 July 2023) was prompted with “please explain the following in simpler terms” along with each institutional PEM to produce revised responses. The readability of both the institutional and GPT-4 revised PEMs was evaluated using validated readability formulas through readable.com. Finally, a board-certified cardiologist reviewed institutional and revised PEMs to assess for changes in accuracy and comprehensiveness.

**Results:** A total of 86 PEMs were analyzed. None of the institutional PEMs met the recommended 6th-grade reading level and had a median Flesch-Kincaid Grade Level (FKGL) of 10.9 (high school freshman; IQR: 9.2, 12.6, p<0.001). In comparison, GPT-4 revised PEMs reduced the median FKGL to 7.8 (seventh grade; IQR: 7.0, 8.8, p<0.001), with 14/86 (16.3%) achieving at least a 6th-grade reading level. All GPT-4 revised PEMs 86/86 (100%) were accurate after review, with 9/86 (10.5%) deemed more comprehensive and 3/86 (3.5%) deemed less comprehensive than the institutional materials.

**Conclusions:** GPT-4 significantly improved the readability of institutional PEMs for CA while maintaining accuracy and, in some cases, enhancing comprehensiveness. These findings underscore the potential of LLMs to bridge health literacy gaps by simplifying complex medical information without compromising content integrity. However, further research is needed to validate our findings and assess patient comprehension, real-world efficacy, and the impact of AI-driven education on clinical outcomes.

## INTRODUCTION

Cardiac amyloidosis (CA) is a progressive, infiltrative, and multi-system disease characterized by the deposition of insoluble misfolded protein aggregates [1]. In the United States, approximately 3,000 new cases of amyloid light chain amyloidosis are diagnosed annually, classifying it as an orphan disease [2, 3]. Globally, it remains rare, with a 20-year prevalence of 51 cases per million [4]. The development of heart failure in patients with CA adds an additional burden, requiring patients to consistently adhere to lifestyle modifications and extensive guideline-based medication regimens. Given the complex challenges associated with CA, enhancing the readability of patient education materials (PEMs) could play a crucial role in improving health literacy and patient care through more effective self-management and increased understanding of treatment [5].

The average American adult reading comprehension level is at the 7th-8th grade proficiency [6]. As a result, the American Medical Association (AMA) recommends that PEMs be written at a 5th-6th grade reading level [7]. Studies have demonstrated that patient education significantly influences health behaviors and there is a connection between low health literacy and poor clinical outcomes in managing cardiovascular disease [8–10]. Despite this, a readability analysis of cardiovascular related PEMs from various online articles and from the AHA website revealed that the mean reading levels were comparable to that of a high school sophomore [11, 12].

Chat Generative Pre-trained Transformer (ChatGPT) is a large language model (LLM) that has rapidly gained traction with high public utilization. [13]. With more patients turning to the internet for health information this novel technology may have significant applications in healthcare [14]. LLMs may offer promising avenues for enriching patient education and simplifying the complex content of PEMs. As ChatGPT’s use and acceptance has expanded, early research primarily focused on assessing its accuracy and reliability. Multiple studies have demonstrated that ChatGPT is proficient across a wide range of medical topics and even making clinical decisions [15–19]. With its accuracy rigorously tested, emerging research is now shifting its attention to evaluating the readability of ChatGPT’s responses.

A recent readability evaluation of online CA PEMs provided by academic medical centers found an average reading level equivalent to that of a high school junior, well above the AMA’s recommendation [20]. Another study also found that while GPT-4 responses regarding CA were primarily accurate and comprehensive, the majority again exceeded the recommended reading levels [18]. Additionally, GPT-4 has demonstrated its ability to improve the readability of PEMs for procedures and conditions such as bariatric surgery, heart failure, uveitis, skull base surgery, and melanoma, highlighting its potential to simplify complex medical information and enhance patient understanding [21–25]. Given the shortage of PEMs for CA and rare diseases that are written at more understandable reading levels [26], improving the readability of these materials may play a critical role in enhancing patient outcomes through improving patient health literacy. We aimed to build on previous research by evaluating the readability of online CA PEMs from leading academic institutions, assessing GPT-4’s ability to improve their readability, and comparing the accuracy and comprehensiveness of the original materials with those revised by GPT-4.

## METHODS

### Institutional Patient Education Materials

We collected 88 PEMs (Supplement A.1) related to CA during July 2023 from the top 10 cardiology institutions listed in the 2022-2023 U.S. News & World Report as “Best Hospitals for Cardiology, Heart & Vascular Surgery.” Institution names were deidentified during analysis. Two PEMs were excluded as they contained multimedia content that could not be processed by GPT-4 at the time, resulting in 86 PEMs analyzed in the study (Figure 1). These frequently asked questions (FAQs) covered various aspects of CA care, including disease overview, causes, types, symptoms, diagnostics, treatments, prognosis, and complications. Duplicate PEMs were retained, as slight content variations between institutions were relevant given the study’s primary focus on readability.

**Figure 1.**
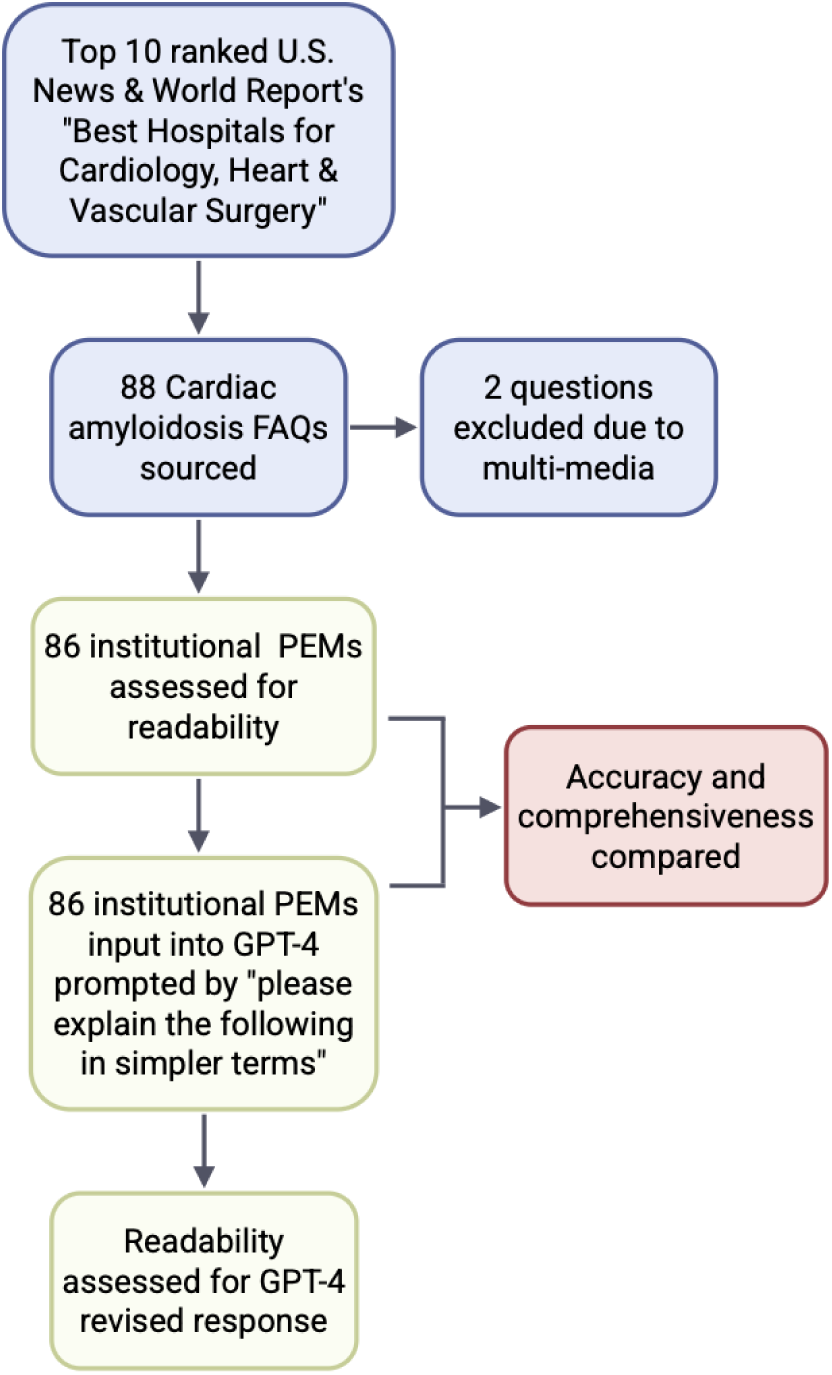
Data collection process for this study including PEM curation, GPT-4 revised PEM generation, and subsequent assessment of readability, accuracy, and comprehensiveness of PEMs. Figure created using biorender.com.

### GPT-4 Response Generation

Each institutional PEM was entered into GPT-4 (version updated 20 July 2023) using the prompt “Please explain the following in simpler terms.” The resulting responses were copied into an Excel file without modification. GPT-4 was accessed via the OpenAI website with default settings, and each PEM was separately processed in a “new chat” session to ensure independent responses. Non-text institutional materials, such as images or video, were excluded since GPT-4 could not process these multimedia components at the time of data collection.

### Readability Assessment

We assessed the readability of both institutional PEMs and GPT-4 revised PEMs using several validated and commonly used formulas: the Flesch Reading Ease Score (FRE) [27], Flesch-Kincaid Grade Level (FKGL) [28], Gunning Fog Index (GFI) [29], Coleman-Liau Index (CLI) [30], SMOG Index [31], and Automated Readability Index (ARI) [32]. FRE is scored from 0 to 100, with higher values indicating greater readability. The other metrics estimate the U.S. grade level required for comprehension; for example, a score of 12 corresponds to a 12th-grade level, while lower scores indicate that lower reading comprehension is needed to understand the text. These formulas derive their scores from parameters such as average sentence length and word complexity. Similar to other studies, readability scores for all formulas were generated by uploading institutional and GPT-4 revised PEMs to readable.com (Supplement A.2) to assess the ease of comprehension [23, 33].

### Accuracy and Comprehensiveness

Accuracy and comprehensiveness of the GPT-4 revised PEMs were assessed by a board-certified cardiologist specializing in cardiac amyloidosis at a tertiary academic medical center. The cardiologist used the following grading scale to compare each revised PEM with its corresponding institutional version: “Compared to the institutional PEM the GPT-4 revised PEM is:” 1) less accurate, 2) equally accurate, or 3) more accurate and “Compared to the institutional PEM, the GPT-4 revised PEM is:” 1) less comprehensive, 2) equally comprehensive, or 3) more comprehensive.

### Statistical Analysis

Descriptive statistics are reported as medians with interquartile ranges (IQR). The Mann-Whitney U test was used to compare readability metrics between institutional and GPT-4 revised PEMs. A sub-analysis evaluated the proportion of PEMs meeting the AMA’s recommended 6th-grade reading level. Statistical analyses were performed using Microsoft Excel version 16.94 and IBM SPSS version 29.

### Ethical Considerations

This observational study did not include patient data and therefore did not require de-identification or data protection measures.

## RESULTS

The GPT-4 revised PEMs demonstrated significant improvements in readability across all metrics compared to the original institutional PEMs, with all results achieving a p-value of <0.001 (Table 1, Figure 2). The Flesch-Kincaid Grade Level showed a notable reduction, with the institutional PEMs having a median of 10.9 (IQR: 9.2, 12.6; p<0.001) compared to 7.8 in the GPT-4 revised PEMs (IQR: 7.0, 8.8; p<0.001), achieving a reading level of a 7th to 8th-grade student and approaching the AMA’s recommended 5th-6th grade level. The absolute reduction in the median Flesch-Kincaid Grade Level was 3.1 grade levels after GPT-4 revision. The Flesch Reading Ease Score improved from a median of 41.0 (IQR: 28.5, 49.8; p<0.001) in the institutional PEMs to 62.5 (IQR: 58.3, 69.4; p<0.001) in the GPT-4 revised versions. The Automated Readability Index also improved, decreasing from 10.3 (IQR: 8.3, 11.6; p<0.001) in the institutional PEMs to 8.0 (IQR: 6.9, 9.0; p<0.001) in the GPT-4 revisions. Additionally, the Gunning Fog Index, Coleman-Liau Index, and SMOG Index for the GPT-4 revised PEMs had medians of 9.2 (IQR: 8.1, 10.4; p<0.001), 9.9 (IQR: 8.8, 11.1; p<0.001), and 10.4 (IQR: 9.2, 11.0; p<0.001), respectively, compared to the higher medians in the institutional PEMs of 12.7 (IQR: 11.0, 14.7; p<0.001), 12.8 (IQR: 11.6, 14.7; p<0.001), and 12.7 (IQR: 11.4, 14.0; p<0.001), respectively. Additionally, following GPT-4 revision, 14/86 (16.3%) of PEMs achieved the AMA’s recommended 5th-6th grade reading level, compared to 0% of the original institutional PEMs (Table 2).

**Figure 2.**
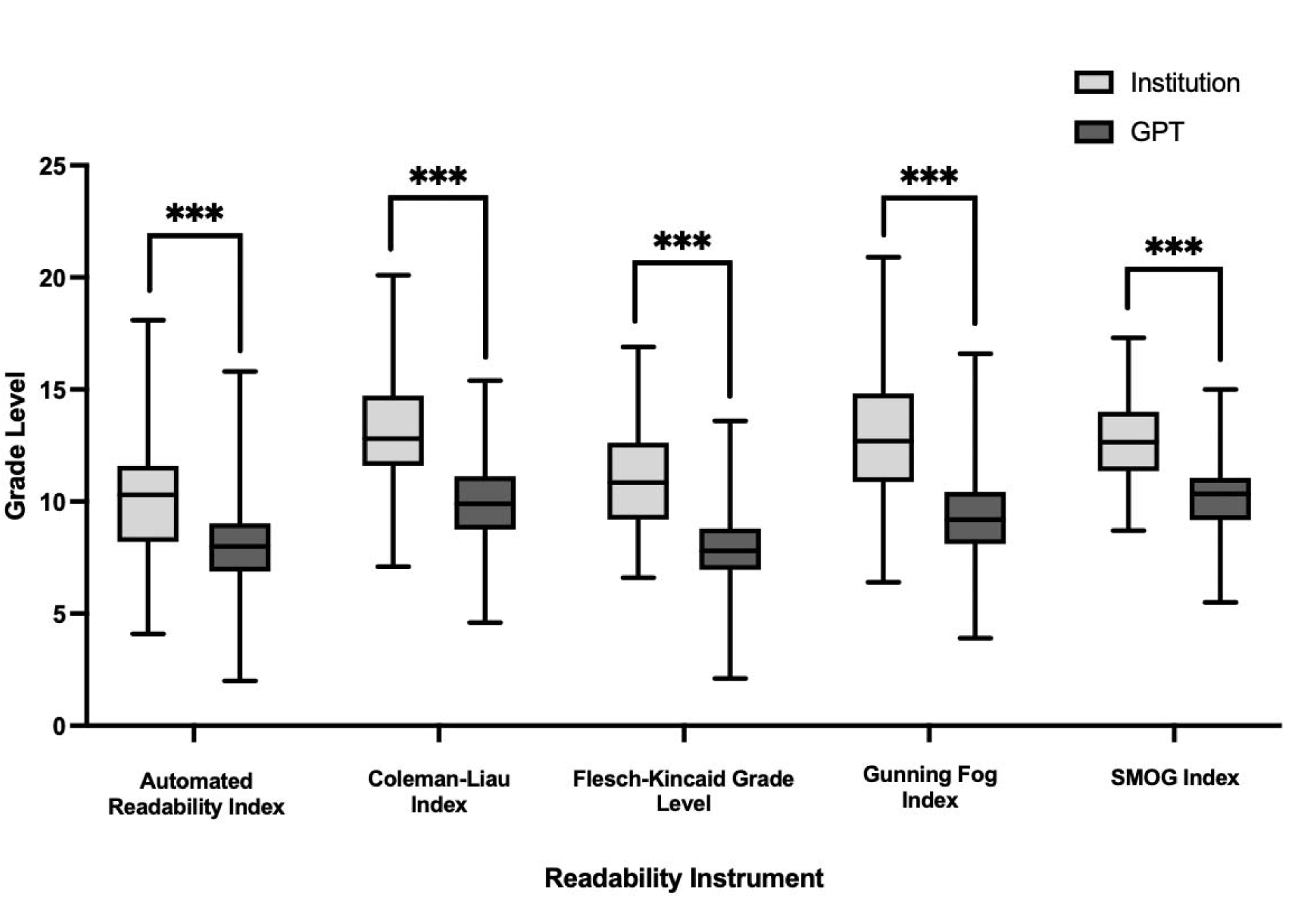
Box and whiskers plot of median readability scores across five metrics for institutional and GPT-4 revised PEMs.

**Table 1.** Readability comparison of institutional and GPT-4 revised PEMs.

|  | Institutional |  |  | GPT-4 |  |  | p-value |
| --- | --- | --- | --- | --- | --- | --- | --- |
| Readability Metrics | Median | IQR (25th, 75th) |  | Median | IQR (25th, 75th) |  |  |
| Flesch Reading Ease score | 41.0 | 28.5 | 49.8 | 62.5 | 58.3 | 69.4 | <.001 |
| Flesch-Kincaid Grade Level | 10.9 | 9.2 | 12.6 | 7.8 | 7.0 | 8.8 | <.001 |
| Gunning Fog Index | 12.7 | 11.0 | 14.7 | 9.2 | 8.1 | 10.4 | <.001 |
| Coleman-Liau index | 12.8 | 11.6 | 14.7 | 9.9 | 8.8 | 11.1 | <.001 |
| SMOG index | 12.7 | 11.4 | 14.0 | 10.4 | 9.2 | 11.0 | <.001 |
| Automated readability index | 10.3 | 8.3 | 11.6 | 8.0 | 6.9 | 9.0 | <.001 |

**Table 2.** Comparison of institutional and GPT-4 revised PEMs meeting the AMA’s recommended 5th-6th grade reading level. Values represent the number and percentage of PEMs at or below this level.

|  | **<=6 Reading Level** | **>=6 Reading Level** | **Percent Meeting**<br>**AMA**<br>**Recommendation** |
| --- | --- | --- | --- |
| **Institutional Flesch-Kincaid Grade Level** | 0.0 | 86.0 | 0.0% |
| **GPT Flesch-Kincaid Grade Level** | 14.0 | 74.0 | 16.3% |

After review by a board-certified cardiologist with expertise in cardiac amyloidosis, the accuracy and comprehensiveness of the GPT-4 revised PEMs were evaluated (Table 3). All GPT-4 revised PEMs 86/86 (100.0%) were deemed equally accurate compared to the original institutional PEMs, with no instances of reduced or increased accuracy. In terms of comprehensiveness, the majority of GPT-4 revised PEMs 74/86 (86.0%) were rated as equally comprehensive as their institutional counterparts, while 9/86 (10.5%) were considered more comprehensive and 3/86 (3.5%) were rated as less comprehensive.

**Table 3.** Assessment of the accuracy and comprehensiveness of GPT-4 revised PEMs compared to institutional PEMs after evaluation by board-certified cardiologist.

| Scoring | Accuracy | Comprehensiveness |
| --- | --- | --- |
| N = 86 responses | Number of responses (%) |  |
| **Less** | 0 (0.0) | 3 (3.5) |
| **Equal** | 86 (100) | 74 (86.0) |
| **More** | 0 (0) | 9 (10.5) |

In cases where the GPT-revised PEMs were deemed more comprehensive, the revisions often featured expanded explanations, clearer organization, and added context to enhance patient understanding. This is exemplified in the explanation to the question of “how amyloidosis is diagnosed?” While the institutional PEM simply listed diagnostic tests such as blood work, imaging studies, tissue biopsy, bone marrow biopsy, nerve conduction study, and genetic testing, the GPT-4 revised PEM offered detailed descriptions of each. It explained blood tests as a method to check for abnormal substances, clarified how imaging studies like echocardiograms visualize heart function, and elaborated on tissue biopsies, specifically heart tissue biopsy, to confirm amyloid deposits. These additions resulted in a clearer understanding of the diagnostic process and the purpose behind each test, contributing to the increased comprehensiveness of the revised material.

## DISCUSSION

Large language models have the potential to enhance patient education materials by delivering information in a conversational format through a user-friendly interface. In this study, we evaluated GPT-4’s ability to improve the readability of institutional PEMs for CA. Our findings demonstrated significant improvements in the readability of institutional PEMs after GPT-4 revision across all readability formulas. After review by a board-certified cardiologist, GPT-4 revised PEMs preserved content accuracy and, in some cases, improved comprehensiveness. Our study provides early evidence showing the potential utility of LLMs for enhancing PEMs with the ultimate future goal of improving patient health literacy and clinical outcomes.

Previous studies have highlighted ChatGPT’s extensive medical knowledge, particularly in cardiology, where it has demonstrated proficiency in acute coronary syndrome, hyperlipidemia, heart failure, atrial fibrillation, and has even aided cardiology team decisions for treating aortic stenosis [15, 34–38]. ChatGPT has also demonstrated the ability to interpret image-based prompts related to Advanced Cardiovascular Life Support (ACLS) and Basic Life Support (BLS) guidelines [39] and has shown promise in detecting CA in patients undergoing transcatheter aortic valve replacement [40]. Beyond cardiology, ChatGPT has exhibited understanding of gastrointestinal diseases, such as cirrhosis, hepatocellular carcinoma, bariatric surgery, and even answered cirrhosis-related questions in non-English languages [21, 41–43]. Despite the rarity of CA, GPT-4 has also been shown to provide accurate and comprehensive answers related to multi-systemic questions related to cardiology, gastroenterology, and neurology [18]. While accurate and comprehensive, ChatGPT’s responses in this study had low readability at baseline, averaging a grade level of 15.5, which is comparable to a college junior. Similar trends were observed in other cardiology topics such as cardiac catheterization and atrial fibrillation [44, 45], as well as in conditions like hypothyroidism during pregnancy and retinal surgical treatments, where accuracy was preserved but readability exceeded recommended levels [46, 47]. Our study demonstrates GPT-4’s ability to adapt and improve the readability of its outputs when prompted, generate responses closer to the AMA-recommended reading levels all while maintaining comparable accuracy and comprehensiveness.

Recent studies have shown that ChatGPT shows promise in improving the readability of online PEMs, although targeted prompting was required in some cases. Preliminary studies found that GPT-3.5 improved aortic stenosis PEMs to a 6th-7th grade level, close to the AMA’s recommended 5th-6th-grade level, but only when specific prompting was used [48]. However, for bariatric surgery, GPT-4 significantly improved PEM readability to meet the AMA’s guidelines without targeted prompting [21]. Similarly, a recent study demonstrated that GPT-4 improved the readability of heart failure PEMs, further highlighting its potential to simplify complex cardiovascular health information [22]. Building on these findings, our study demonstrates that GPT-4 can enhance the readability of CA PEMs by reducing the median reading level from 10.9 to 7.8. Notably, all institutional PEMs did not meet the AMA’s recommended reading level, but after GPT-4 revision, 16.3% reached the AMA’s recommendation without specific prompting. These results underscore the potential of ChatGPT as an interactive platform to improve patient access to PEMs, enhance health literacy, and facilitate better understanding of CA.

### Limitations and Ethical Considerations

Despite their natural conversational interface and ease of use that facilitate engaging, real-time dialogue on complex medical topics, LLMs have limitations that should be addressed before widespread implementation in healthcare. An important limitation is “artificial hallucinations,” where LLMs confidently present inaccurate information that could mislead clinicians or patients. Solutions to this critical limitation are under active investigation. One approach to address this is explainable artificial intelligence (AI), where models provide their source of information. One such example is OpenEvidence, which automatically cites published sources and guidelines, potentially improving accuracy and reliability [49]. Another concern is data privacy. While technology companies like OpenAI allow users to opt out of data collection, cybersecurity breaches and data leaks also pose potential threats, underscoring the need for robust security measures that do not collect patient data to protect privacy.

The implementation of LLMs also raises ethical concerns regarding bias, accessibility, and its potential to reinforce patient care disparities. Research has shown that GPT-4 reinforces stereotypes related to gender, ethnicity, and clinical presentations, contributing to bias in medical decision-making. ChatGPT-3.5 and 4 have been found to perpetuate outdated race-based medical practices, such as factoring African American race into GFR and lung capacity calculations [50]. ChatGPT-3.5 has also exhibited gender and racial bias in acute coronary syndrome management when prompted with patients from female, African American, or Hispanic backgrounds [51]. Beyond bias, accessibility remains a concern, as individuals from lower socioeconomic backgrounds often have limited internet access [52]. This issue is compounded by limitations in some LLMs, including restricted features, query limits, and access to its most advanced versions behind paywalls [53], possibly creating barriers for socioeconomically disadvantaged patients. Furthermore, LLMs convenient and decisive responses may cause some patients to rely on it, rather than seeking care from a medical professional and thus potentially delaying diagnosis and treatment. These challenges underscore the need for clinician oversight to ensure LLM development prioritizes equity, safety, and reduces patient care disparities.

While our study offers valuable insights into the use of ChatGPT-based models for generating patient education materials, several limitations and opportunities for improvement warrant further discussion. The sample was limited to 86 Q&A pairs from a select group of institutions, which may not capture the full spectrum of patient education needs. Moreover, accuracy and comprehensiveness were evaluated by a single expert reviewer, introducing potential subjective bias and limiting generalizability. Additionally, the study did not assess patient understanding or clinical outcomes following GPT-4 implementation, nor did it include multimedia questions, as the version of ChatGPT (GPT-4) used in this study lacked this capability at the time of data collection. Future research should broaden the scope to include a larger, more diverse set of PEMs across various conditions and institutions, incorporate multiple reviewers to reduce bias, and explore the use of a more updated GPT version with enhanced multimedia support. Furthermore, developing custom ChatGPT versions tailored specifically for medical applications holds promise for generating even more accurate and applicable PEMs. Finally, future studies could randomize patients to receive either ChatGPT-generated or traditional education materials, then measure clinical outcomes to evaluate the real-world effectiveness of GPT-supported patient education.

## Conclusion

GPT-4 significantly improved the readability of institutional PEMs for CA while maintaining accuracy and, in some cases, enhancing comprehensiveness. These findings underscore the potential of LLMs to bridge health literacy gaps by simplifying complex medical information without compromising content integrity. However, further research is needed to assess patient comprehension, real-world efficacy, and the impact of AI-driven education on clinical outcomes. Future studies should explore the integration of LLMs into patient education strategies, mitigate potential biases, and evaluate their role in reducing disparities in health literacy. Rigorous validation, clinician oversight, and ethical considerations will be essential to ensuring the safe and equitable implementation of AI in patient-centered care.

## Abbreviations

AI: Artificial Intelligence
AMA: American Medical Association
ARI: Automated Readability index
ChatGPT: Chat Generative Pre-trained Transformer
CLI: Coleman-Liau Index
FKGL: Flesch-Kincaid Grade Level
FRE: Flesch Reading Ease score
GFI: Gunning Fog Index
IQR: Interquartile Range
LLM: Large Language Model
PEMs: Patient Education Materials
SMOG: Simple Measure of Gobbledygook

## Acknowledgements

N/A.

## References

1. Kittleson, M.M., et al., Cardiac Amyloidosis: Evolving Diagnosis and Management: A Scientific Statement From the American Heart Association. Circulation, 2020. 142(1): p. e7–e22.

2. Maurer, M.S., et al., Addressing Common Questions Encountered in the Diagnosis and Management of Cardiac Amyloidosis. Circulation, 2017. 135(14): p. 1357–1377.

3. Rossi, M., et al., Re-Definition of the Epidemiology of Cardiac Amyloidosis. Biomedicines, 2022. 10(7).

4. Kumar, N., et al., Global epidemiology of amyloid light-chain amyloidosis. Orphanet Journal of Rare Diseases, 2022. 17(1): p. 278.

5. Berkman, N.D., et al., Low health literacy and health outcomes: an updated systematic review. Ann Intern Med, 2011. 155(2): p. 97–107.

6. Statistics, N.C.f.E. The NCES Fast Facts Tool Provides Quick Answers to Many Education Questions (National Center for Education Statistics). 2019; Available from: https://nces.ed.gov/fastfacts/display.asp?id=69.

7. Weiss, B.D., Health literacy: a manual for clinicians. 2003: American Medical Association Foundation and American Medical Association.

8. Magnani, J.W., et al., Health Literacy and Cardiovascular Disease: Fundamental Relevance to Primary and Secondary Prevention: A Scientific Statement From the American Heart Association. Circulation, 2018. 138(2): p. e48–e74.

9. Safeer, R.S., C.E. Cooke, and J. Keenan, The impact of health literacy on cardiovascular disease. Vasc Health Risk Manag, 2006. 2(4): p. 457–64.

10. van Schaik, T.M., et al., Cardiovascular disease risk and secondary prevention of cardiovascular disease among patients with low health literacy. Neth Heart J, 2017. 25(7-8): p. 446–454.

11. Ayyaswami, V., et al., A Readability Analysis of Online Cardiovascular Disease-Related Health Education Materials. Health Lit Res Pract, 2019. 3(2): p. e74–e80.

12. Kapoor, K., et al., Health Literacy: Readability of ACC/AHA Online Patient Education Material. Cardiology, 2017. 138(1): p. 36–40.

13. Center, P.R., Americans’ use of ChatGPT is ticking up, but few trust its election information. March 26, 2024.

14. Center, P.R. The Social Life of Health Information. June 11th, 2009; Available from: https://www.pewresearch.org/internet/2009/06/11/the-social-life-of-health-information/.

15. King, R.C., et al., Appropriateness of ChatGPT in Answering Heart Failure Related Questions. Heart Lung Circ, 2024.

16. Rao, A., et al., Assessing the Utility of ChatGPT Throughout the Entire Clinical Workflow: Development and Usability Study. J Med Internet Res, 2023. 25: p. e48659.

17. Sarraju, A., et al., Appropriateness of Cardiovascular Disease Prevention Recommendations Obtained From a Popular Online Chat-Based Artificial Intelligence Model. JAMA, 2023. 329(10): p. 842–844.

18. King, R.C., et al., A Multidisciplinary Assessment of ChatGPT’s Knowledge of Amyloidosis: Observational Study. JMIR Cardio, 2024. 8: p. e53421.

19. Ayers, J.W., et al., Comparing Physician and Artificial Intelligence Chatbot Responses to Patient Questions Posted to a Public Social Media Forum. JAMA Internal Medicine, 2023. 183(6): p. 589–596.

20. Byer, S. and U. Grewal, Abstract 4148214: Missing the Point: Readability Analysis of Online Patient Information for Cardiac Amyloidosis. Circulation, 2024. 150(Suppl_1): p. A4148214–A4148214.

21. Srinivasan, N., et al., Large language models and bariatric surgery patient education: a comparative readability analysis of GPT-3.5, GPT-4, Bard, and online institutional resources. Surg Endosc, 2024. 38(5): p. 2522–2532.

22. King, R., et al., Abstract 4145543: ChatGPT-4 Improves Readability of Institutional Heart Failure Patient Education Materials. Circulation, 2024. 150(Suppl_1): p. A4145543-A4145543.

23. Mohammadi, S.S., et al., Evaluation of the Appropriateness and Readability of ChatGPT-4 Responses to Patient Queries on Uveitis. Ophthalmology Science, 2025. 5(1): p. 100594.

24. Warn, M., et al., Assessing the Readability, Reliability, and Quality of AI-Modified and Generated Patient Education Materials for Endoscopic Skull Base Surgery. Am J Rhinol Allergy, 2024. 38(6): p. 396–402.

25. Diniz-Freitas, M., et al., Assessing the accuracy and readability of ChatGPT-4 and Gemini in answering oral cancer queries—an exploratory study. Exploration of Digital Health Technologies, 2024. 2(6): p. 334–345.

26. Falcão, M., et al., A Community-Based Participatory Framework to Co-Develop Patient Education Materials (PEMs) for Rare Diseases: A Model Transferable across Diseases. Int J Environ Res Public Health, 2023. 20(2).

27. Flesch, R. Guide to academic writing. University of Canterbury School of Business and Economics. 2016; Available from: https://web.archive.org/web/20160712094308/http://www.mang.canterbury.ac.nz/writing_guide/writing/flesch.shtml.

28. Kincaid J, F.R., Rogers R, Chissom B Derivation of New Readability Formulas (Automated Readability Index, Fog Count and Flesch Reading Ease Formula) for Navy Enlisted Personnel. 1975.

29. Gunning, R., The Fog Index After Twenty Years. Journal of Business Communication, 1969. 6(2): p. 3–13.

30. Coleman, M. and T.L. Liau, A computer readability formula designed for machine scoring. Journal of Applied Psychology, 1975. 60(2): p. 283–284.

31. Mc Laughlin, G.H., SMOG Grading-a New Readability Formula. Journal of Reading, 1969. 12(8): p. 639–646.

32. Smith, E.A., et al., Automated readability index. AMRL-TR-66-220. 1967, Wright-Patterson Air Force Base, Ohio: Aerospace Medical Research Laboratories, Aerospace Medical Division, Air Force Systems Command. iii, 14 pages.

33. Reddy, R.V., et al., Assessing the quality and readability of online content on shock wave therapy for erectile dysfunction. Andrologia, 2022. 54(11): p. e14607.

34. Gurbuz, D.C. and E. Varis, Is ChatGPT knowledgeable of acute coronary syndromes and pertinent European Society of Cardiology Guidelines? Minerva Cardiol Angiol, 2024. 72(3): p. 299–303.

35. Lee, T.J., et al., Evaluating ChatGPT-3.5 and ChatGPT-4.0 Responses on Hyperlipidemia for Patient Education. Cureus, 2024. 16(5): p. e61067.

36. Lee, T.J., et al., Abstract 17100: Evaluating Chatgpt Responses on Atrial Fibrillation for Patient Education. Circulation, 2023. 148(Suppl_1): p. A17100-A17100.

37. Altamimi, I., et al., The scientific knowledge of three large language models in cardiology: multiple-choice questions examination-based performance. Ann Med Surg (Lond), 2024. 86(6): p. 3261–3266.

38. Salihu, A., et al., A study of ChatGPT in facilitating Heart Team decisions on severe aortic stenosis. EuroIntervention, 2024. 20(8): p. e496–e503.

39. King, R.C., et al., GPT-4V passes the BLS and ACLS examinations: An analysis of GPT-4V’s image recognition capabilities. Resuscitation, 2024. 195: p. 110106.

40. Pereyra Pietri, M., et al., The prognostic value of artificial intelligence to predict cardiac amyloidosis in patients with severe aortic stenosis undergoing transcatheter aortic valve replacement. European Heart Journal - Digital Health, 2024. 5(3): p. 295–302.

41. Yeo, Y.H., et al., Assessing the performance of ChatGPT in answering questions regarding cirrhosis and hepatocellular carcinoma. Clin Mol Hepatol, 2023. 29(3): p. 721–732.

42. Samaan, J.S., et al., Assessing the Accuracy of Responses by the Language Model ChatGPT to Questions Regarding Bariatric Surgery. Obes Surg, 2023. 33(6): p. 1790–1796.

43. Yeo, Y.H., et al., GPT-4 outperforms ChatGPT in answering non-English questions related to cirrhosis. medRxiv, 2023: p. 2023.05.04.23289482.

44. Behers, B.J., et al., Assessing the Readability of Patient Education Materials on Cardiac Catheterization From Artificial Intelligence Chatbots: An Observational Cross-Sectional Study. Cureus, 2024. 16(7): p. e63865.

45. Lee, T.J., et al., Evaluating ChatGPT Responses on Atrial Fibrillation for Patient Education. Cureus, 2024. 16(6): p. e61680.

46. Momenaei, B., et al., Appropriateness and Readability of ChatGPT-4-Generated Responses for Surgical Treatment of Retinal Diseases. Ophthalmol Retina, 2023. 7(10): p. 862–868.

47. Onder, C.E., et al., Evaluation of the reliability and readability of ChatGPT-4 responses regarding hypothyroidism during pregnancy. Sci Rep, 2024. 14(1): p. 243.

48. Rouhi, A.D., et al., Can Artificial Intelligence Improve the Readability of Patient Education Materials on Aortic Stenosis? A Pilot Study. Cardiol Ther, 2024. 13(1): p. 137–147.

49. Wu, V. and J. Casauay, OpenEvidence.

50. Omiye, J.A., et al., Large language models propagate race-based medicine. NPJ Digit Med, 2023. 6(1): p. 195.

51. Zhang, A., et al., ChatGPT Exhibits Gender and Racial Biases in Acute Coronary Syndrome Management. medRxiv, 2023: p. 2023.11.14.23298525.

52. Wang, X., et al., ChatGPT: promise and challenges for deployment in low- and middle-income countries. Lancet Reg Health West Pac, 2023. 41: p. 100905.

53. Center, O.H., What Is CHATGPT plus?

